# Effects of gender-affirming hormone therapy on gray matter density, microstructure and monoamine oxidase A levels in transgender subjects

**DOI:** 10.1101/2022.04.29.22274480

**Authors:** PA Handschuh, MB Reed, M Murgaš, C Vraka, U Kaufmann, L Nics, M Klöbl, M Ozenil, ME Konadu, EM Patronas, B Spurny-Dworak, A Hahn, M Hacker, M Spies, P Baldinger-Melich, GS Kranz, R Lanzenberger

## Abstract

MAO-A catalyzes the oxidative degradation of monoamines and is thus implicated in sex-specific neuroplastic processes that influence gray matter (GM) density (GMD) and microstructure (GMM). Given the exact monitoring of plasma hormone levels and sex steroid intake, transgender individuals undergoing gender-affirming hormone therapy (GHT) represent a valuable cohort to potentially investigate sex steroid-induced changes of GM and concomitant MAO-A density. Here, we investigated the effects of long-term GHT over a median time period of 4.5 months on GMD and GMM as well as MAO-A distribution volume. To this end, 20 cisgender women, 11 cisgender men, 20 transgender women and 10 transgender men underwent two MRI scans in a longitudinal design. PET scans using [^11^C]harmine were performed before each MRI session in a subset of 35 individuals. GM changes determined by diffusion weighted imaging (DWI) metrics for GMM and voxel based morphometry (VBM) for GMD were estimated using repeated measures ANOVA. Regions showing significant changes of both GMM and GMD were used for the subsequent analysis of MAO-A density. These involved the fusiform gyrus, rolandic operculum, inferior occipital cortex, middle and anterior cingulum, bilateral insula, cerebellum and the lingual gyrus (post-hoc tests: p_FWE+Bonferroni_ < 0.025). In terms of MAO-A distribution volume, no significant effects were found. Additionally, the sexual desire inventory (SDI) was applied to assess GHT-induced changes in sexual desire, showing an increase of SDI scores among transmen. Changes in the GMD of the bilateral insula showed a moderate correlation to SDI scores (rho = −0.62, p_Bonferroni_ = 0.047). The present results are indicative of a reliable influence of gender-affirming hormone therapy on 1) GMD and GMM following an interregional pattern and 2) sexual desire specifically among transmen.

**Highlights:**

- Gender-affirming hormone therapy led to significant changes in gray matter density and microstructure in various brain regions
- Gray matter changes found after gender-affirming hormone therapy were not reflected by monoamone oxidase A density changes in the brain.
- Masculinizing gender-affirming hormone therapy resulted in increased sexual desire in transgender men.

## Introduction

As pointed out in several reviews, meta- and mega-analyses that have been published over the last decades, a multitude of studies on sex-specific differences in brain structure point towards significant effects of sex steroids in terms of global, inter- and intra-regional brain morphology (1–5). On average, female brains are smaller than male ones (6). Significant differences between male and female brains were also found regarding gray (GM) and white matter (WM) volumes (7, 8). Nevertheless, some studies failed to detect such differences in GM volume on a global scale, despite adjustment for total brain size (9). Both region-of-interest (ROI)-based approaches as well as voxel-wise analyses allow for the examination and classification of highly specific cortical and subcortical regions, shedding light on a vast number of potential regional and/or local sex differences (3–5). However, a closer look to the current evidence reveals some inconsistent results, questioning the theory of a sexually dimorphic brain and pointing towards a lack of features found reliably at one end of the continuum between typically female and male brains (10).

Even though previous literature has reported conflicting results, it is known that sex hormones are centrally involved in the biological mechanisms that drive sex-specific differences in brain structure. Hence, ongoing research focusing on the interaction of cycling sex steroid levels with components of the central nervous system is essential. Sex steroids are known to have both transcriptional as well as non-genomic effects (11–13), modifying the brain throughout the lifespan (14–17). Thus, distributional differences in sex hormone receptors across the brain (18–20) as well as sex-specific differences in axonal characteristics (21), neuroplasticity (22) and neurogenesis (23) can be found. Additionally, sex steroids are known to directly interact with various neurotransmitters (24–26) and their metabolism, e.g., by modulating the activity of cerebral MAO-A activity. This has already been shown in a number of animal models (27–30) and selected human studies (31), however *in vivo* imaging data acquired via positron emission tomography (PET) remains scarce (32–34) and studies investigating the direct effects of exogenously administered sex hormones on cerebral MAO-A are specifically underrepresented (35). A further gap in the literature that needs to be addressed is given by studies investigating the effect of *MAOA* gene polymorphisms that were shown to interact with GM changes – despite knowledge on the influence of MAOA VNTR polymorphism on brain structure volumes (36), cortical thickness (37) and gray matter (38), the association of MAO-A protein expression as measured in human *in vivo* studies and GM is insufficiently investigated. Taking into account the relevance of MAO-A density and subsequent neurotransmitter metabolism on neuroplastic effects (39–41) and thus GM characteristics in the brain, the link between MAO-A distribution and GM needs to be explored with multimodal imaging techniques. Taking into account the additional effect of sex steroids on both MAO-A density (32–34) as well as GM (7, 8), sex-specific interrelations of MAO-A density and GM might be of interest.

Individuals with gender dysphoria (GD) receiving gender-affirming hormone therapy (GHT) represent a unique cohort to investigate the direct effects of sex hormones on human brain structure. As described in the 5^th^ edition of the Diagnostic and Statistical Manual of Mental Disorders (DSM-V), GD is defined as an incongruence between the biological sex assigned at birth and the experienced gender later in life (42). GHT basically involves the long-term administration of an estrogen and a type of androgen blocker in transgender women (TW), whereas the masculinization of transgender men (TM) can be supported by testosterone treatment (43). As summarized by Kranz and colleagues, structural changes of the brain were observed after GHT in a wide range of studies (44), with effects of testosterone treatment on total brain volume (45, 46), GM and WM density (47–49) as well as cortical thickness, showing controversial results in the latter (47, 50). In terms of estrogen and antiandrogen treatment, a decrease in cortical thickness (47), total brain (45) as well as total GM and WM volume (46) was shown. Changes in diffusivity as it was found after sex steroid treatment both in TM and TW might be traced back to changes in the organizational properties of neural fibers (e.g., axon diameter, fiber density, myelin staining) (44). On a molecular level, high-dose testosterone treatment was shown to influence the distribution volume (V_T_) of MAO-A in transgender individuals (33). The structural and molecular effects of sex steroids on the brain are also reflected by behavioralchanges, e.g., in terms of sexual behavior. In a cross-sectional multi-centered transgender study, Garz et al. could show that masculinizing GHT was associated with increased sexual desire in transmen (51). This is in line with the findings of Defreyne et al. (52), who reported a long-lasting increase of sexual desire as measured by the sexual desire inventory (SDI) in TM after GHT, whereas in TW, multiple SDI subscores decreased during the first three months of GHT.

In this longitudinal study, we aimed to investigate changes to brain morphology and microstructure and further explore the corresponding effects on MAO-A binding in transgender individuals after long-term GHT. To reveal alterations in GM density (GMD) and microstructure (GMM) via MRI, voxel-based morphometry (VBM) and diffusion weighted imaging (DWI) were applied. In a subset of participants, PET imaging using [^11^C]harmine was conducted to uncover the effects of sex steroid treatment on cerebral MAO-A density and to elucidate potential interrelations between hormonally induced GM changes and alterations in MAO-A V_T_. In addition, we aimed to reveal the effect of GHT on sexual desire in both transmen and transwomen.

## Methods

This study was performed according to the Declaration of Helsinki, taking into account all current revisions and good scientific practice guidelines of the Medical University of Vienna. The study protocol was approved by the Ethics Committee of the Medical University of Vienna (EC number 1104/2015) and registered at clinicaltrials.gov (NCT02715232).

### Participants

Twenty cisgender women (CW), 11 cisgender men (CM), 20 TW and 10 TM men were enrolled in the study. Transgender subjects were recruited at the Unit for Gender Identity Disorder at the Department of Obstetrics & Gynecology, Division of Gynecologic Endocrinology and Reproductive Medicine, Medical University of Vienna, Austria. Control subjects were recruited via designated message boards at the Medical University of Vienna and at other universities in Vienna as well as via social media platforms. CW and CM were age and education level matched to TW and TM, respectively. Importantly, for transgender individuals, inclusion criteria comprised a diagnosis of gender dysphoria as defined in the Diagnostic and Statistical Manual for Mental Disorders), version 5 (DSM-5: 302.85) or the International Classification of Diseases, version 10 (ICD-10: F64.1). Moreover, the absence of severe psychiatric, neurologic or internal comorbidities was ensured in case of study enrollment based on medical history, physical examination, electrocardiogram, laboratory screening and structural clinical interview (SCID) for DSM-V. All subjects gave written informed consent to take part in this study and received financial compensation for their participation. Subjects were excluded in case of major neurological or internal illnesses, pregnancy, current substance abuse (excluding smoking), any kind of pathologic laboratory values, GHT or any other treatment with exogenous sex hormones within 6 months prior to study enrollment, MRI contraindications, current or past substance-related disorder or non-compliance (all subjects), any kind of current or prior psychiatric diagnosis (CW and CM) or severe DSM-V comorbidities (TW and TM) as well as current (TW and TM) or prior (CW and CM) treatment with psychotropic agents.

### Study design

All subjects underwent two MRI scans separated by a median time period of 4.5 months following a longitudinal study design. Each MRI session included structural (T1- and diffusion-weighted), task-based and resting-state MRI as well as MR spectroscopy. For the current analyses, only structural MRI data was used. Thirty-five subjects (9 CW, 8 CM, 7 TW and 11 TM), underwent PET scan(s) that were performed before MRI acquisition using the radiotracer [^11^ C]harmine. After the baseline imaging sessions, transgender subjects received GHT. Plasma hormone levels were assessed via blood draw right before or after each baseline and follow-up imaging session. In addition, participants underwent the sexual desire inventory 2(SDI-2) to assess potential effects of GHT on participants’ interest in sexual activity.

### Medication

The following GHT protocol was applied at the Department of Obstetrics & Gynecology, Division of Gynecologic Endocrinology and Reproductive Medicine, Medical University of Vienna, Austria: TM were treated with either 1000 mg testosterone undecanoat every 8-16 weeks (Nebido 1000 mg / 4 ml i.m.), or up to 50 mg testosterone daily (Testogel 50 mg/day or Testavan 23 mg/pump 1-2 pumps/day or testosterone crème as a magistral formula 12,5 mg/pump 3-4 pumps/day transdermally). If menstruation was still observed, transgender male participants received either lynestrenol (Orgametril 5 mg 2-3 tablets/day p.o.) or desogestrel (Moniq Gynial or Cerazette 0.075 mg/day p.o.). TW received cyproterone acetate daily (Androcur 50 mg/day) and either estradiol transdermally (Estramon 100 μg transdermal patch 2x/week or Estrogel gel 0.75-1.5 mg/day transdermally) or oral estradiol (Estrofem 4 mg/day p.o.). In case of extensive hair loss, an alpha-5-reductase-inhibitor was prescribed (Finasterid 2.5 mg/48h Actavis/Arcana/Aurobindo p.o.). Furthermore, selected participants were treated with GnRH-analogues like triptorelin (decapeptyl 0.1 mg/day subcutaneously).

### Sex steroid monitoring

Levels of luteinizing hormone (LH), follicle-stimulating hormone (FSH), progesterone, estradiol, testosterone, sex hormone binding globulin (SHBG) and dehydroepiandrosterone sulfate were assessed via blood analysis at baseline and after a median time period of 4.5 months involving GHT in transgender subjects and no treatment in cisgender controls.

### Magnetic resonance imaging acquisition

All MRI data were recorded using a Siemens Prisma 3T scanner and a 64-channel head coil. A whole-brain T1-weighted scan was acquired with the following parameters: TE / TR = 2.91 / 2000 ms; inversion time = 900 ms; flip angle = 9°; matrix = 240 x 256, 176 slices; voxel size = 1.0 mm^3^; acquisition time = 7:59 min. Diffusion weighted images contained 64 diffusion encoded images, with a b-value of 800 s/mm^2^ along with 7 non-diffusion (b=0) images (TE / TR=76 / 9400 ms, slice thickness = 1.6 mm, resolution=1.6 x 1.6 mm, matrix size=128 x 128 x 75, flip angle = 90°, acquisition time = 11:54 min). Subjects were instructed to stay awake with eyes open and keep their head as still as possible during the entire scan to minimize movement artifacts.

### Positron emission tomography

Synthesis and quality control of [^11^C]harmine, (7-[^11^C]methoxy-1-methyl-9H-pyrido[3,4-b]indole) were performed as previously published (53). All PET scans were performed using a GE Advance full-ring scanner (General Electric Medical Systems, Milwaukee, WI, USA) at the Department of Biomedical Imaging and Image-guided Therapy, Division of Nuclear Medicine, Medical University of Vienna (54–56). A 5 min transmission scan was done at the beginning of the scan for tissue attenuation correction. Data acquisition started simultaneously with an intravenous bolus injection of [^11^C]harmine (4.6 MBq/kg body weight). Ninety minutes of PET data were acquired in 3D mode and reconstructed using an iterative filtered backprojection algorithm, which was separated into 51 time frames with a spatial resolution of 4.36 mm with a FWHM of 1 cm next to the center of the field of view and reconstructed in 35 transaxial sectional volumes (128 × 128 matrix). Automatic arterial blood sampling was performed during the first 10 min at a rate of 4 ml/min (ALLOGG, Mariefred, Sweden) and manual samples were taken at 5, 6, 7, 8, 10, 20, 40, 60, and 80 min after bolus start (57). A gamma counter previously cross-calibrated to the PET scanner was used to measure radioactivity concentrations in both whole blood and plasma. Furthermore, samples at 6, 7 and 8 min were used for individual cross-calibration between manual and automated blood sampling. Fractions of radioactive metabolites and the parent compound were determined with HPLC as described previously (58).

### MRI data processing

Voxel-based morphometric changes were calculated and processed in the CAT12 toolbox (http://www.neuro.uni-jena.de/cat/, version r1742) in MATLAB (version 9.8) using the longitudinal pipeline with default settings unless otherwise specified. Prior to preprocessing, raw data was visually inspected for potential artifacts. In short, the T1 images from both time points were co-registered, resampled and bias-corrected. Each scan was then skull stripped and segmented into GM, white matter and cerebrospinal fluid, and resulting GMD was further used. Finally, GMD maps were normalized into MNI space and spatially smoothed using an 8-mm Gaussian kernel. Lastly, the CAT12 reports and data were visually controlled for registrations and segmentation errors.

As an estimate for GMM changes, DWI data was processed within the FMRIB software library (FSL, v. 6.0.05) (59) and unring (https://bitbucket.org/reisert/unring), ANTs (https://github.com/ANTsX/ANTs), MRtrix (https://www.mrtrix.org/) according to an optimized pipeline (60). All data was brain-masked, corrected for noise, movement, gradient, Gibbs ringing, bias field and eddy current distortions as well as for outliers in the form of signal dropouts (60). The diffusion tensors were fitted using the “dtifit” command as well as the rotated b-vectors and then smoothed using a 1 mm kernel (60). Mean diffusivity (MD) estimates were calculated and co-registered with the participant- and measurement-specific segmented GMD map. Thereafter, all images were normalized into MNI space and subsequently the GMD segmentation was thresholded and used to mask out non-GM voxels. Finally, the DWI MD estimates were smoothed using an 8 mm Gaussian kernel and masked again.

### PET data processing and MAO-A quantification

MAO-A quantification was carried out using PMOD 3.509 (PMOD Technologies Ltd., Zurich, Switzerland; www.pmod.com). Whole blood data was fitted with the sum of three exponentials from the peak onward, plasma-to-whole blood ratio was fitted with a linear function and the parent fraction was modeled with a Watabe function. The arterial input function (AIF) was obtained by multiplying the fitted whole blood activity, plasma-to-whole blood ratio and the fraction of intact radioligand in plasma. The Logan plot was used to quantify voxel-wise MAO-A total V_T_ with the thalamus as high uptake region. Regional V_T_ were extracted using the activation map created from the overlap between GMD and DWI analyses.

### Statistics

Structural changes determined by the MD DWI metric and GMD were estimated using 4 x 2 (group x time) repeated measures ANOVA (rmANOVA) implemented in SPM12. Interactions between group (TM, TW, CW, CM) by time (before and after min. 4 months of hormone therapy) were examined and further corrected for multiple testing using Gaussian random field theory as implemented in SPM12 and the threshold for significance was set at p ≤ 0.025 family-wise error (FWE)-corrected according to (61) at the cluster-level following P ≤ 0.001 uncorrected at the voxel-level. Further post-hoc comparisons and number of DWI metrics were adjusted again using the Bonferroni method. To account for the individualized GHT treatment, progesterone, testosterone and estradiol were added to both rmANOVA models as covariates. Furthermore, the total intracranial volume was added as a covariate in the VBM model. For both the GMD and DWI models each covariate was standardized, and group mean centered.

To provide a robust identification of hormonally induced gray matter alterations, the different structural changes reflected by both the GMD and DWI analyses were combined. The significant effects from both models were binarized and a conjunction map across both imaging modalities was calculated as an intersection. The remaining brain regions included in this conjunction map were used for the subsequent MAO-A V_T_ analysis.

Interaction effects in MAO-A V_T_ before and after GHT for both cis- and transgender individuals were estimated using a linear mixed model in SPSS 25.0 (SPSS Inc., Chicago, Illinois; www.spss.com). Here, the global model consisted of group, time and ROI (extracted from the conjunction map) as factors and MAO-A V_T_ as the dependent variable. The model was reduced in case of non-significance. Paired t-tests were used to estimate changes in SDI scores before and after GHT for both transmales and transfemales (p_Bonferroni_ < 0.025). Finally, partial correlations between changes in SDI scores to GMD and GMM while correcting for the main influencing hormone were calculated to investigate a possible link between structural and behavioral changes after GHT (p_Bonferroni_ < 0.05). The Bonferroni method was used to correct for multiple comparisons in all tests.

## Results

Demographics and hormone levels are listed in table 1. Fewer TW and matched CM were recruited when compared to TM and CW. Due to irreversible damage to the PET imaging equipment, 35 participants partook in the PET part of the study, where only 17 completed both scans, see table 1 for a detailed breakdown of subgroups. Out of the 35 participants that underwent the PET scans, only 52 scans were suitable for further analysis.

**Table 1:** Demographics of all participants for both DWI + VBM and PET analyses. each divided into their respective subgroups. Age is given from the inclusion date. Hormone levels for the pre-treatment assessment and cisgender participants are provided for comparison only and were not used as covariates in the statistical analyses.

|  | cisgender women |  | cisgender men |  | transgender women |  | transgender men |  |
| --- | --- | --- | --- | --- | --- | --- | --- | --- |
| N(VBM + DWI) | 20 |  | 11 |  | 20 |  | 10 |  |
| Age [years] | 25.3 ± 6.7 |  | 26.5 ± 8.0 |  | 28.6 ± 9.3 |  | 24.75 ± 7.3 |  |
| Completed | M1 | M2 | M1 | M2 | M1 | M2 | M1 | M2 |
| N(PET) | 9 | 3 | 8 | 3 | 7 | 5 | 11 | 6 |
|  | Pre | Post | Pre | Post | Pre | Post | Pre | Post |
| DHEAS (µg/ml) | 3.2 ± 0.9 | 3.2 ± 1.0 | 3.8 ± 1.9 | 2.6 ± 1.3 | 3.9 ± 1.1 | 3.8 ± 1.3 | 3.2 ± 1.9 | 3.3 ± 1.9 |
| Estradiol (pg/ml) | 81.2 ± 56.7 | 112.1 ± 79.5 | 22.7 ± 10.3 | 22.9 ± 9.0 | 35.7 ± 16.6 | 290.0 ± 299.1 | 111.2 ± 74.9 | 59.3 ± 50.3 |
| FSH (µU/ml) | 4.9 ± 1.4 | 4.9 ± 2.8 | 3.8 ± 1.6 | 3.8 ± 1.5 | 3.3 ± 1.7 | 0.2 ± 0.1 | 4.5 ± 2.5 | 5.6 ± 3.6 |
| LH (µU/ml) | 9.8 ± 6.4 | 9.5 ± 6.9 | 5.8 ± 1.9 | 6.5 ± 2.3 | 6.5 ± 2.9 | 0.1 ± 0.1 | 9.6 ± 5.9 | 8.6 ± 8.8 |
| Progesterone (ng/ml) | 1.3 ± 1.3 | 4.9 ± 5.6 | 0.4 ± 0.3 | 0.3 ± 0.2 | 0.3 ± 0.2 | 0.8 ± 0.9 | 4.9 ± 6.4 | 0.3 ± 0.2 |
| SHBG (nmol/L) | 67.4 ± 28.5 | 69.6 ± 27.6 | 45.3 ± 25.0 | 43.2 ± 19.3 | 45.7 ± 21.5 | 70.2 ± 39.4 | 65.3 ± 36.3 | 36.3 ± 15.1 |
| Testosterone (ng/ml) | 0.4 ± 0.2 | 0.3 ± 0.1 | 5.6 ± 2.1 | 5.0 ± 1.2 | 5.7 ± 1.7 | 0.3 ± 0.2 | 0.4 ± 0.2 | 4.3 ± 2.1 |

### Structural and MAO-A changes after GHT

For both the VBM and DWI analyses, significant 2-way (time-by-group) interaction effects were found. More widespread structural changes after a median of 4.5 months GHT using VBM were discovered, when compared to MD, see figure 1a + b. Combining the results from VBM and DWI analyses showed an overlap of changes in the fusiform gyrus, rolandic operculum, inferior occipital cortex, middle and anterior cingulum and insula, cerebellum and the lingual gyrus (figure 1c). Post-hoc tests revealed changes in GMD, see figure 2 and also for MD, figure 3, (p _FWE + Bonferroni_ < 0.025). An increase in GMD was found in the fusiform gyrus, cerebellum and lingual gyrus after GHT for transmen compared to both control groups. Transwomen contrastingly showed a decrease in GMD after GHT in the rolandic operculum, insula, posterior cingulum, cerebellum and lingual gyrus when compared to both cismales and cisfemales.

**Figure 1:**
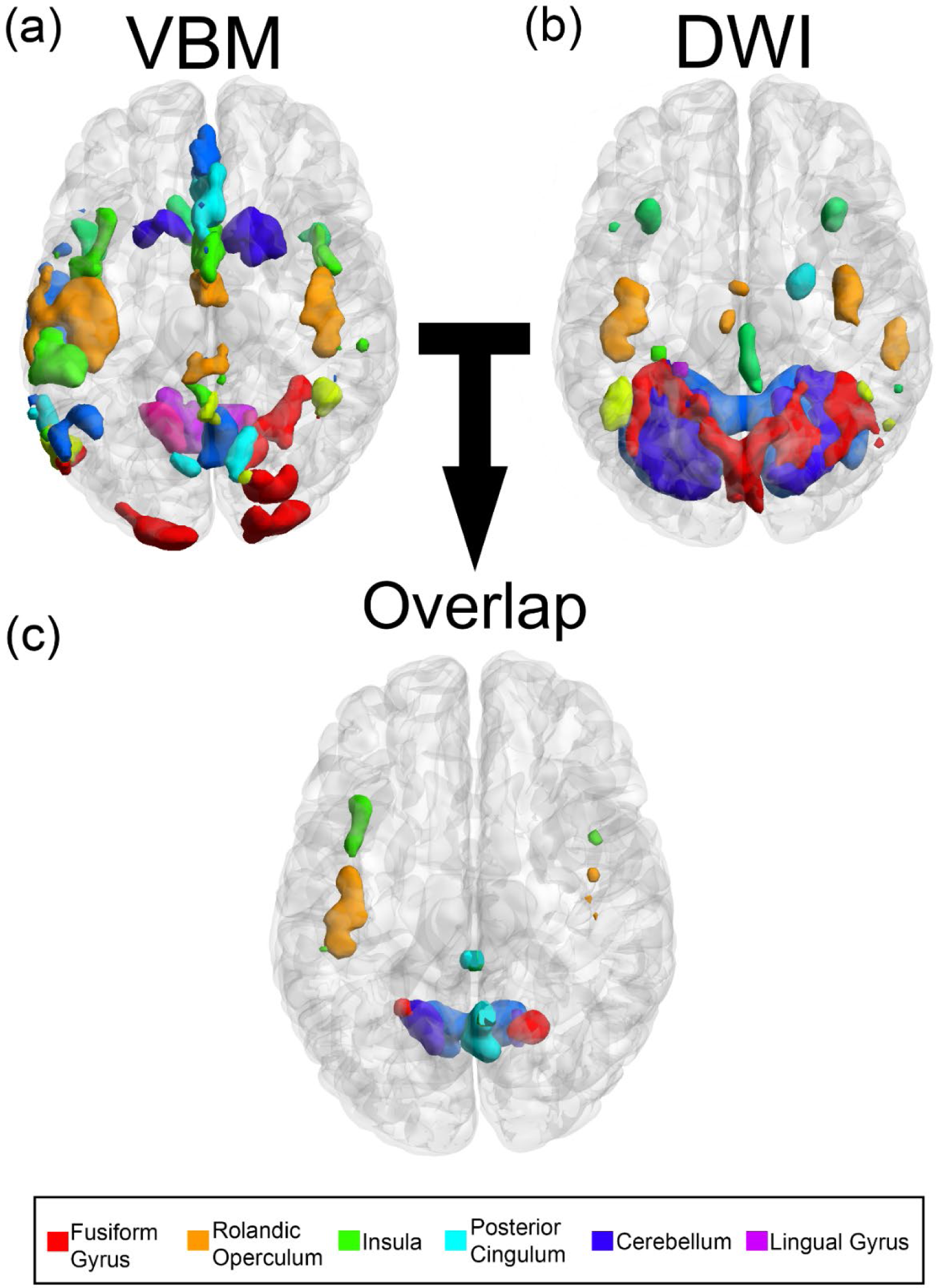
Structural changes after gender-affirming hormone therapy. (a) Shows the significant changes derived from the voxel based morphometry (VBM) analysis, whereas (b) shows the mean diffusivity changes in gray matter for all transgender and cisgender groups after a median of 4.5 months of gender-affirming hormone therapy. (c) Indicates the overlapping regions between the VBM and DWI analyses.

**Figure 2:**
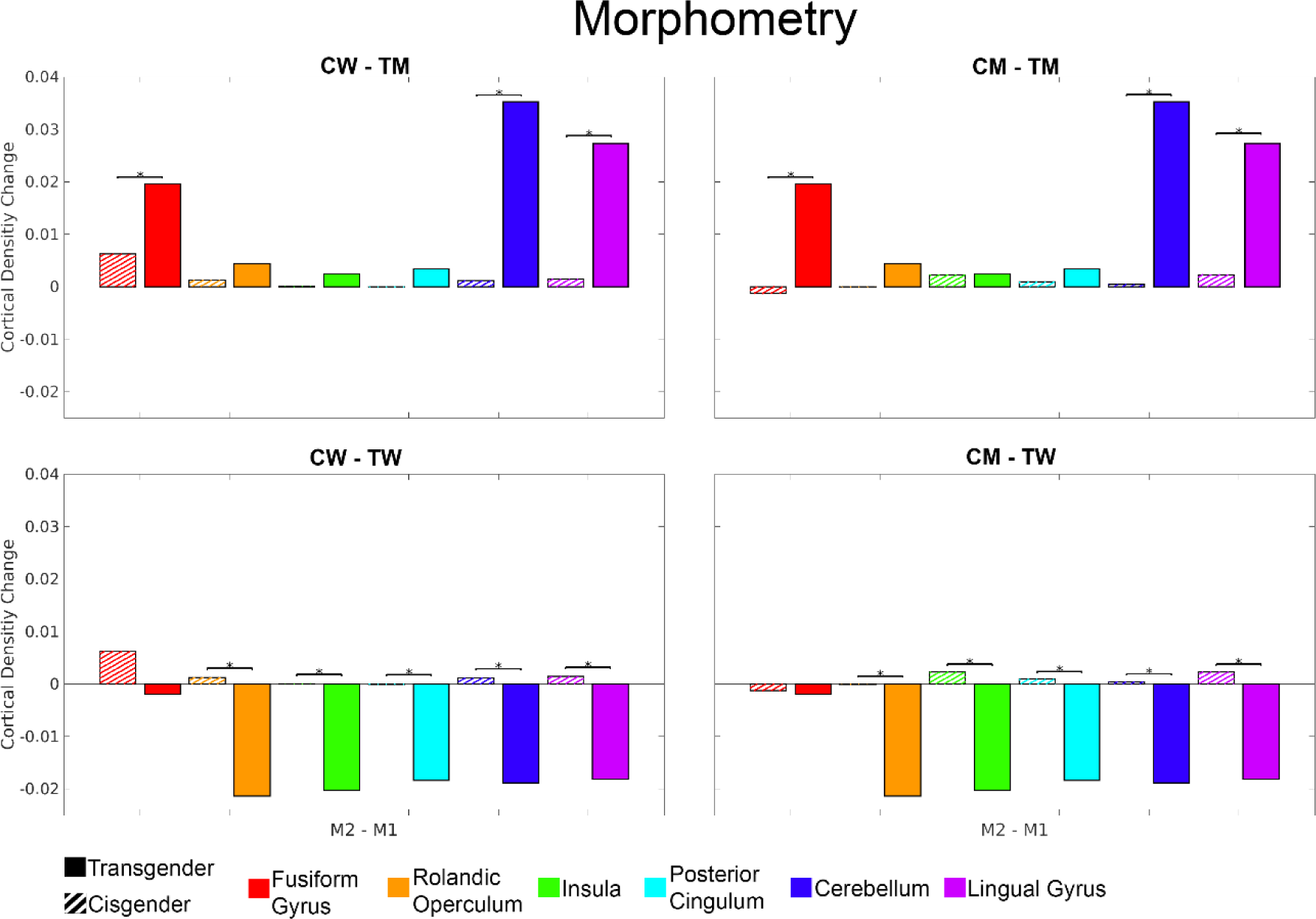
Voxel based morphometry regional changes. Each bar represents the change in gray matter density (GMD) after 4 months of gender-affirming hormone therapy (GHT), where the hatched bars represent the cisgender groups of male (CM) and female controls (CW) and the solid bars the transgender groups, transmale (TM) and transfemale (TW). Here widespread regional interactions can be found in the TW group when compared to both CM and CW, whereas TM showed less regional interactions, however the significant interactions showed a greater magnitude of change. An increase in GMD was found TM, whereas a decrease was found in TW after a median of 4.5 months of GHT when compared to both CM and CW. Significant group-by-time interactions are denoted with a *.

**Figure 3:**
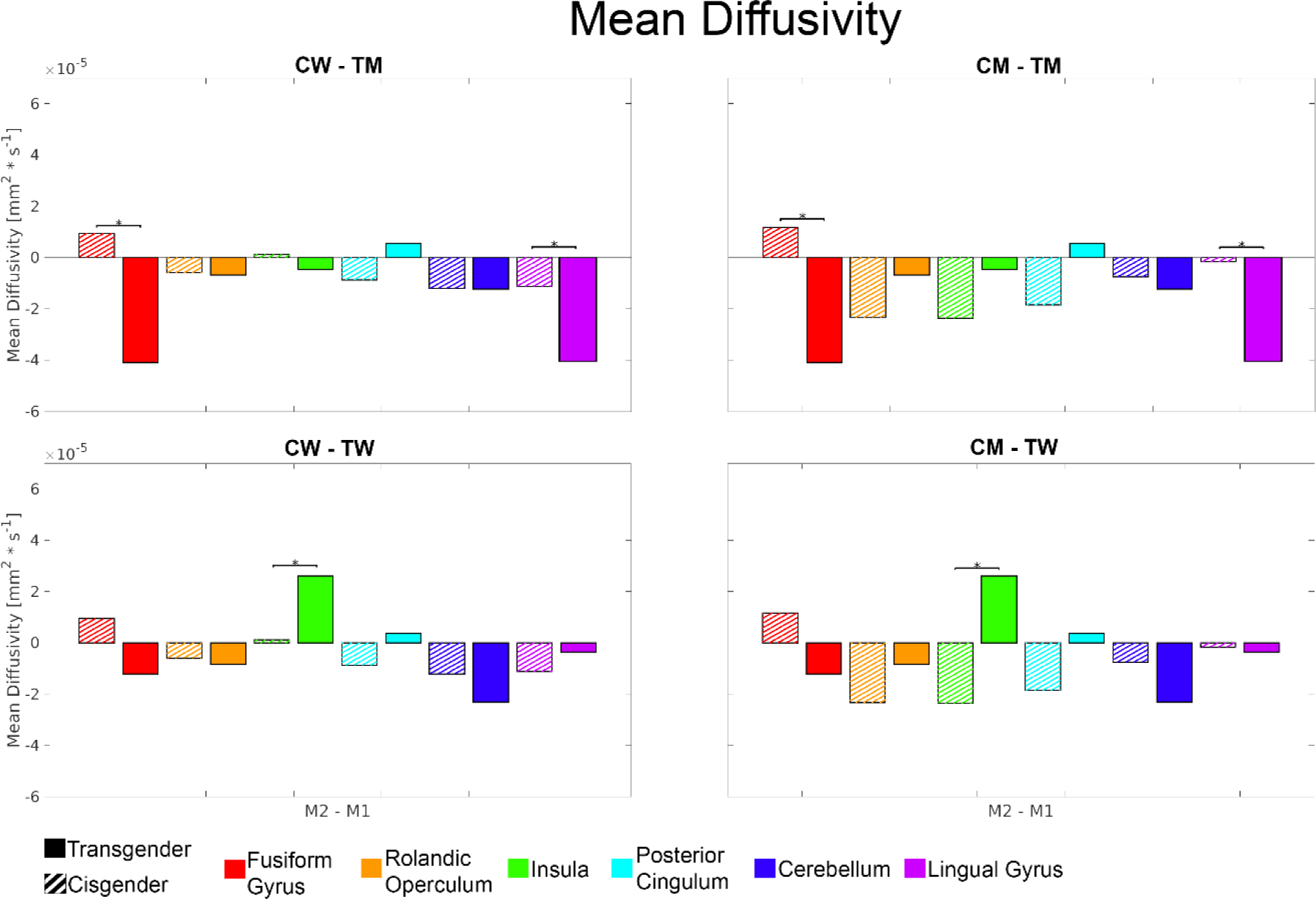
Mean diffusivity regional changes. Each bar represents the average diffusivity change reflective of microstructural alterations after 4 months of gender-affirming hormone therapy, where the hatched bars represent the cisgender groups of male (CM) and female controls (CW) and the solid bars the transgender groups, transmale (TM) and transfemale (TW). Here, changes a decrease in both TM vs CW or CM and also TW vs CM were found. Significant group-by-time interactions are denoted with a *.

The opposite interaction effects were found in the MD of the fusiform and lingual gyrus for transmales compared to both cis control groups. An increase of MD in the insula was also found for transwomen after GHT when compared to both control groups.

When investigating regional changes in MAO-A V_T_, no interaction and no main effects were significant.

### Behavioral changes after GHT

An increase in the overall SDI scale (sum score) “desire with (t = 5.84, p_Bonferroni_ < 0.001) and without interactions” (t = 3.4, p_Bonferroni_ = 0.01) was found for TM after GHT. Furthermore, a moderate negative correlation between changes in the SDI sum score of desire with interaction and GMD in the bilateral insula after GHT for the TM group was found (rho = −0.62, p_Bonferroni_ = 0.047). While other regions and sum score of desire without interaction also showed moderate correlations, no others survived the correction for multiple comparisons. Moreover, no significant correlations between GMM and any SDI sum scores were found, p > 0.12.

## Discussion

The present study aimed to investigate potential changes to GM and MAO-A density induced by GHT over a median time period of 4.5 months in subjects with GD. To reveal robust changes in both GD, morphometric differences in GMD and microstructural alterations as reflects by GMM were combined and subsequently used for assessment of MAO-A V_T_. Significant changes in GMD and GMM after GHT that will be discussed in the following section were found in nine distinct brain regions. Across these regions, no significant changes of MAO-A V_T_ were found. In an additional analysis we probed the effect of GHT on sexual desire as measured by the SDI and found an increase in the SDI sum scores “desire with” and “without interactions” for TM after GHT.

### Structural changes after GHT

Significant 2-way (time-by-group) interaction effects induced by GHT, as reflected by both the VBM and DWI analyses, were found in the fusiform gyrus, rolandic operculum, inferior occipital cortex, middle and anterior cingulum, bilateral insula, cerebellum as well as the lingual gyrus. As discussed in the following section, our findings are in line with prior studies reporting significant changes in GMD and GMM. Moreover, in comparison to cross-sectional data (62–66), our longitudinal design allows for more accurate interpretation.

#### Fusiform gyrus

As various other regions presenting significant changes to GMD and GMM reported here, the fusiform gyrus is implicated in own-body perception in the context of self (49, 67) and is known to show different functional connectivity patterns in GD when compared to cisgender controls (49). These functionally different patterns might be reflected by structural differences as well. Here, we found a GHT-induced increase of GMD and decrease of MD (as a marker for GMM) in the fusiform gyrus of TM, potentially indicating anticatabolic effects of testosterone (44, 68).

#### Lingual gyrus, bilateral insula and posterior cingulum

After androgenization of TM, Zubiaurre-Elorza and colleagues found a significant increase of cortical thickness in multiple brain regions, e.g., the left lingual gyrus (47). This is in line with our finding, however in the present study, GMD of the lingual gyrus increased and MD decreased bilaterally across TM after high-dose testosterone treatment. Of note, the lingual gyrus is centrally involved in processing visual information and thus visuospatial functioning. As men, on average, perform better than women in visuospatial functioning, and testosterone has been suggested to directly influence visuospatial functioning both in men (69) as well as women (70), it could be hypothesized that advantages in this skill could partially be traced back to an interrelation of higher GMD and and lower and the influence of high-dose testosterone treatment. In addition, we found a significant decrease in GMD across the insula as well as the posterior cingulum of TW when compared to CW and CM. The insula belongs to the salience network (71), which plays a central role for interoceptive processes and own-body perception (67). Differences in functional connectivity were further shown in language and affective processing (72, 73) as well as in various neuropsychiatric conditions (74, 75). However, sex-based differences between cisgender men and women regarding the insula were not only shown in functional MRI, but also in structural analyses (3) and investigations of the GABAergic system in humans (76). Thus, changes to GMD might be seen as a marker for microstructural remodeling induced by estrogen and anti-androgen therapy, as it is needed for multifactorial adaptations that take place throughout GHT (49, 77). Our findings are in line with prior reports on decreased GMD, higher MD reflecting microstructural changes, and widespread cortical thinning in anti-androgen and estrogen treatment across TW (46, 47).

### Changes to MAO-A distribution

In view of sexually dimorphic characteristics of multiple neuropsychiatric conditions, better understanding of how sex steroids interact with neurotransmitters on a molecular level, e.g., as reflected by MAO-A density, is of major interest. In the present study, none of the subgroups showed significant changes to MAO-A V_T_ in any of the brain regions that were found to be different in terms of GMD and GMM after GHT. In contrast, in a prior analysis of MAO-A density after GHT performed by our group, we found statistically significant reductions of MAO-A V_T_ in six of twelve previously defined regions of interest in TM after GHT for at least 4 months. These regions included the middle frontal cortex (−10%), anterior cingulate cortex (−9%), medial cingulate cortex (−10.5%), insula (−8%), amygdala (−9%) and hippocampus (−8.5%, all p<0.05)) (33). Of note, in the study by Kranz et al. regional MAOA V_T_ was estimated based on *a priori* defined regions-of-interest (ROIs) from a modified AAL atlas (33, 78), whereas here, MAO-A analysis was performed in regions that showed significant GMD and GMM changes after GHT.

### GHT effects on sexual desire

The effect of GHT on sexual desire found here corroborates the findings of previous observations reporting increased sexual desire in TM after androgen treatment (51, 52, 79–82). Even though Defreyne et al. found that some sexual desire changes observed over the course of GHT might diminish over time, elevated total, dyadic, and solitary SDI scores in TM remained stable on the long-term (52). This is in line with our finding of elevated sexual desire in TM after a median time period of 4.5 months of GHT. Despite the effects quantified 4.5 months after GHT initiation, no further follow-up was performed in the present study. Therefore, a decrease of sexual desire after a longer therapeutic period cannot be ruled out completely. However, a follow-up period of 12 months has been performed by van Dijk and colleagues who reported the most pronounced changes in sexual desire across TM within the first 3 months of treatment which remained largely stable even after 12 months (81, 83). The increase in sexual desire among TM might be traced back to testosterone treatment per se (84) and higher androgen levels in general (85). However, many other factors could have influenced sexual drive, e.g., more satisfactory sexual interactions and relationships due to transition or improvement in general well-being after GHT (82).

Interestingly, partial correlation analyses between changes in SDI scores and GMD/GMM revealed a significant negative correlation between changes in the SDI sum score of desire with interaction and GMD in the bilateral insula of TM individuals. This finding suggests a possible connection between structural GM changes in the insula and changes in sexual desire after masculinizing GHT. Among many other functions, the insula is essential for the integration of external sensory stimuli via various neuronal pathways (86). Activation of the bilateral insula during sexual arousal and desire has been shown in several fMRI studies (87–89), indicating a link between insular activation and the processing of sexual stimuli. Although there have been multiple fMRI studies in this area, this is, to the best of our knowledge, the first study to link structural GM changes after long-term masculinizing GHT with changes in sexual desire. Moreover, the significance of a negative (rather than positive) correlation between structural GM changes and changes of sexual desire (found in subjects treated with testosterone) has yet to be evaluated. Nevertheless, the finding points to an influence of the hormone on both sexual desire and structural remodeling within cortical regions such as the insula.

Of note, also the neurotransmitter dopamine plays a key role in insular activation while processing sexual stimuli (90). Thus, the presence of mesolimbic dopaminergic projections reaching the insula and contributing to the processes taking place during sexual arousal and sexual desire should be considered in future neuroimaging studies in this field (91).

### Limitations

Some limitations of the present study need to be addressed. First, subgroups were not sufficiently balanced due to recruitment difficulties, especially in case of subjects with gender dysphoria. Second, the subgroup that underwent PET imaging comprised only 17 individuals due to irreparable damages to the PET equipment. Therefore, it cannot be ruled out that potential GHT-induced changes in MAO-A V_T_ both on a global as could not be detected in the present analyses.

## Conclusion

Long-term GHT seems to have a considerable impact on GMD and GMM in the brain of transgender individuals. Specific effects of either androgenizing or feminizing sex steroids must be taken into account in most regions, however, in selected structures both types of GHT affected GMD and GMM in the same way, pointing towards mechanisms that are induced by GHT irrespective of whether feminizing or androgenizing steroids are used. The combined approach in analyzing structural MRI data has the potential to shed light on changes of GM as reflected by both morphometric as well as microstructural analyses after GHT in a longitudinal design and in comparison to cisgender individuals. Nevertheless, larger sample sizes are needed to detect reliable associations between GM and potential MAO-A density changes induced by GHT. In terms of sexual desire, our findings are consistent with the literature and supports evidence from previous observations of increased sexual desire in TM after masculinizing GHT.

## Acknowledgments

This research was funded in whole, or in part, by the Austrian Science Fund (FWF) [Grant number KLI 504, PI: Rupert Lanzenberger]. M. Murgaš is funded by the Austrian Science Fund (FWF) [Grant number DOC 33-B27, Supervisor R. Lanzenberger]. MB. Reed and M. Klöbl are recipients of a DOC fellowship of the Austrian Academy of Sciences at the Department of Psychiatry and Psychotherapy, Medical University of Vienna. This project was performed with the support of the Medical Imaging Cluster of the Medical University of Vienna, and by the grant „Interdisciplinary translational brain research cluster (ITHC) with highfield MR” from the Federal Ministry of Science, Research and Economy (BMWFW), Austria. We thank the graduated team members and the diploma students of the Neuroimaging Lab (NIL, headed by R. Lanzenberger) as well as the clinical colleagues from the Department of Psychiatry and Psychotherapy of the Medical University of Vienna for clinical and/or administrative support. In particular, we would like to thank R. Seiger (MRI) and L. Rischka (PET) for technical and methodological support and V. Ritter for administrative help. We would also like to thank N. Berroterán-Infante, T. Balber, C. Philippe for assistance with radioligand synthesis, K. Rebhan for radioligand metabolite processing, I. Leitinger, H. Ibeschitz, V. Weiss for assistance with PET measurements, and V. Pichler, M. Mitterhauser, T. Traub-Weidinger and S. Kasper for project supervision.

## CRediT authorship contribution statement

**Patricia A Handschuh**; conceptualization, investigation, visualization, writing – original draft, **Murray B Reed**; conceptualization, methodology, software, formal analysis, data curation, visualization, writing – original draft, **Matej Murgaš**; software, data curation, review & editing, **Crysoula Vraka**; review & editing, **Ulrike Kaufmann**; review & editing, investigation, **Lukas Nics**; review & editing, **Marius Ozenil**; review & editing, **Dr. Melisande E. Konadu**; investigation, review & editing, **Manfred Klöbl**; investigation, data curation, writing – review & editing, **Eva-Maria Patronas**; review & editing, **Benjamin Spurny-Dworak**; review & editing, **Andreas Hahn**; software, supervision, writing – review & editing, **Marcus Hacker**; review & editing, **Marie Spies**; conceptualization, investigation, writing – review & editing, **Pia Baldinger-Melich**; review & editing, investigation, **Georg S. Kranz**; conceptualization, writing – review & editing, **Rupert Lanzenberger**; project administration, funding acquisition, supervision

## Conflict of Interest

With relevance to this work there is no conflict of interest to declare. R. Lanzenberger received travel grants and/or conference speaker honoraria within the last three years from Bruker BioSpin MR, Heel, and support from Siemens Healthcare regarding clinical research using PET/MR. He is a shareholder of the start-up company BM Health GmbH since 2019. G.S. Kranz declares that he received conference speaker honorarium from Roche, AOP Orphan and Pfizer. P. Handschuh received authorship honoraria from MedMedia Verlag and a travel grant from Angelini Pharma. The other authors report no conflict of interest.

## Data availability statement

Due to data protection laws processed data is available from the authors upon reasonable request. Please contact with any questions or requests.

## Notes

### Clinical Trial

NCT02715232

### Author Declarations

The Ethics Committee of the Medical University of Vienna gave ethical approval for this work. (EC number 1104/2015).

### Summary of Updates

Questionnaire added to manuscript; Revision of results and discussion;

